# Pregnancy-related complications and associated factors among women attending antenatal care at a specialised maternal and child health national referral hospital, in Uganda

**DOI:** 10.1101/2022.07.29.22278187

**Authors:** Bridget Nagawa Tamale, Douglas Bulafu, John Bosco Isunju, Atuko Vicky Jamu, James Natweta Baguma, Arnold Tigaiza, Aisha Nalugya, Rogers Musitwa, Richard K. Mugambe, Tonny Ssekamatte, David Lubogo

## Abstract

**Background:** Although pregnancy and child birth-related complications remain a global public health concern, little is known about their prevalence and associated factors. Our study assessed pregnancy-related complications and associated factors among women attending antenatal care at a specialized maternal and child health national referral hospital in Uganda.

**Methodology:** A cross-sectional study was conducted among 285 pregnant women. Data were collected using the KoboCollect mobile application, and analysed using STATA 14. A modified poisson regression model was used for inferential statistics.

**Findings:** Out of the 285 women, 27.4% (78/285) had pregnancy-related complications. The most reported complications were anaemia, 10.9% (31/285); eclampsia, 8.1% (23/285); and still births, 4.9% (14/285). Having a higher gravidity of 4-6 (APR= 0.32, 95% CI: 0.17-0.57) and of more than 7 (APR= 0.32, 95% CI: 0.14-0.73) was negatively associated with pregnancy-related complications. Factors positively associated with pregnancy-related complications included; late first ANC (APR=1.85, 95% CI: 1.17-2.92), parity of ≥ 3 (APR= 3.69, 95% CI: 1.50-9.08) and induced abortion prior to current pregnancy (APR= 1.64, 95% CI: (1.08-2.47).

**Conclusion:** The prevalence of pregnancy-related complications was very high. Anaemia, eclampsia and still births were the most prevalent. A late first ANC, gravidity higher than 4, parity higher than 3, and history of an induced abortion prior to the current pregnancy were associated with having pregnancy-related complications. Interventions aimed at reducing maternal morbidity and mortality should aim at promoting early ANC attendance, and increasing access to safe abortion and family planning services.

## Introduction

Pregnancy and child birth-related complications remain a global public health concern (1). More than 810 women die every day as a result of pregnancy and childbirth related complications (2). Recent evidence indicates that more than 211 deaths occur per 100,000 live births worldwide, 94% of which happen in developing countries (2, 3). With more than 500 maternal deaths per 100,000 live births, Sub Saharan Africa (SSA) accounts for half of the world’s maternal deaths (2). Over the years, Uganda has registered an improvement in maternal health indicators (4, 5), however, this has not been satisfactory. The pregnancy-related mortality ratio (368 deaths per 100,000 live births) and maternal mortality ratio (336 deaths per 100,000 live births) remain high (4).

Nearly three quarters of maternal deaths are due to haemorrhage, unsafe abortion, infections such as sepsis, complications from delivery, and high blood pressure (6, 7). Pregnancy-related complications not only lead to maternal death but also miscarriages, preterm labour or premature rupture of membranes, premature birth, stillbirth, low birth weight, macrosomia, birth defects, and infant morbidity or death (1). Majority of these complications are preventable, and may be diagnosed and managed during antenatal attendance (2, 8, 9).

Antenatal care (ANC) provides an opportunity for the detection and identification of pregnancy-related complications as well as prevention and management of pregnancy-related or concurrent diseases, health education and health promotion (10). It also provides an opportunity to identify women and girls at increased risk of developing complications during labour and delivery, thus ensuring referral for appropriate care (11). Identification and treatment of pregnancy-related complications reduces the maternal mortality ratio, thus contributing to the thirteen targets for Sustainable Development Goal (SDG) 3 which aims to ensure healthy lives and promote well-being for all at all ages (12). SDG 3 targets a reduction in global maternal mortality ratio to 70 per 100,000 live births, neonatal mortality to 12 per 1,000 live births, and under-5 mortality to 25 per 1,000 live births (12).

In order to end preventable maternal deaths, in 2016, the World Health Organization (WHO) developed guidelines on ANC for a positive pregnancy experience, with the aim of streamlining the management of specific pregnancy-related complications (13). These guidelines recommend an early ultrasound scan (before 24 weeks of gestation) for pregnant women to estimate gestational age, improve detection of fetal anomalies and multiple pregnancies, and to reduce induction of labour for post-term pregnancy (13).

WHO guidelines on ANC for a positive pregnancy experience also recommend antibiotics for asymptomatic bacteriuria for the prevention of persistent bacteriuria, preterm birth and low birth weight, antibiotic prophylaxis for the prevention of recurrent urinary tract infections (UTIs), tetanus toxoid vaccination for the prevention of neonatal mortality from tetanus, and intermittent preventive treatment for malaria to reduce the risk of anaemia (13, 14). In Uganda, the goal oriented antenatal care protocol recommends a minimum of 8 ANC visits in an uncomplicated pregnancy (15). The first contact happens in first trimester (0 – 12 weeks), second and third contact occurs during the second trimester (>13 – 28 weeks), while the 4^th^ to 8^th^ contacts occur in the third trimester (29-40 weeks) (15, 16). These visits are used to identify multiple pregnancy and fetal abnormalities, danger signs of pregnancy induced hypertension and any other danger signs, and to provide appropriate preventive and treatment interventions (15, 16).

Both the WHO guidelines on ANC for a positive pregnancy experience, and the Uganda goal oriented antenatal care protocol provide an opportunity for the identification of pregnancy-related complications. However, there is limited evidence of the prevalence of pregnancy-related complications among women attending ANC in Uganda. This study established pregnancy-related complications and associated factors among women attending ANC at a specialised maternal and child health national referral hospital in Uganda.

## Materials and methods

### Study design, setting and population

A cross-sectional hospital-based study employing quantitative data collection methods was conducted among pregnant women attending ANC at Kawempe national referral hospital in December 2021. Kawempe national referral hospital is a specialised maternal and child health national referral hospital in Uganda, located in Kawempe division, one of the five administrative units of Kampala Capital City Authority. It is located approximately 5.5 kilometres, by road, north of Mulago National Referral Hospital, along the Kampala-Gulu highway. It serves neighbouring communities, and referrals from the lower health facilities around Kampala, Wakiso, Mukono and other parts of the country. The hospital runs ANC clinics three times in a week with an average daily attendance of between 200 to 300 pregnant women. Standard ANC package including prenatal ultrasonography is provided to all mothers, and high-risk mothers are offered special investigations such as genetic testing or biophysical profiling (17).

### Sample size and sampling procedures

The sample size was estimated using the Kish Leslie formula (18). An estimated prevalence (p) of 21.5% adopted from a similar study conducted in Bangladesh (19), a 5% margin of error, standard normal deviate at 95% confidence (1.96) and non-response rate of 10% were considered. This yielded a sample size of 285 respondents. Systematic sampling was used in the selection of study participants. First, a list of pregnant women who were attending ANC that day was obtained from the hospital register. Thereafter, every even-numbered participant on the list for each day was selected until a total of 285 women was obtained.

### Data collection technique and tools

Research assistants (RAs) collected data mainly through face-to-face interviews. Interviews were conducted using a structured questionnaire preloaded on Kobo Collect, a mobile data collection software. The Kobo Collect app was preinstalled on android-based mobile phones and data were collected using the devices. The questionnaire elicited data on the socio-demographic characteristics of the pregnant women, obstetric characteristics, knowledge on ANC and pregnancy-related complications. RAs obtained informed consent from each participant and each interview lasted between 20 to 30 minutes.

### Study variables and measurement

The dependent variable for this study was pregnancy-related complications in current pregnancy. In order to assess this variable, respondents were asked whether they had experienced any complications during their current pregnancy. This was coded as “Yes” if a study respondent had experienced a complication, and “No” if she hadn’t. A checklist with outlined complications was used as a guide when probing for pregnancy-related complications among respondents. In this study, pregnancy-related complications were defined as health problems that occurred during pregnancy, which included those affecting the mother’s health, baby’s health or both. The independent variables for this included socio-demographic characteristics such as, age, respondent’s education level, husband’s education level, marital status, respondent’s employment status and partner’s employment status; obstetric characteristics such as parity, gravidity, and ever had induced abortion; and ANC factors including time of first ANC visit, and knowledge on when to start ANC. Knowledge on when to start ANC was assessed as “Yes” if a study respondent mention period between 6-8 weeks into pregnancy, and “No” if she mentioned otherwise. Gravidity was defined as the sum of all pregnancies, including all live births and pregnancies that were terminated at less than6 months or did not result in a live birth. Parity was defined as pregnancies that resulted in the delivery at greater than6 months gestation, of either a live birth or a stillbirth. Late ANC attendance was defined as beyond 20 weeks which is regarded as late ANC attendance according to the Ministry of Health (MOH), Uganda (20).

### Data management and analysis

Data collection and entry was done using Kobo collect and uploaded daily onto the server. Data were downloaded into an excel sheet and then transferred to STATA 14 for data cleaning and statistical analysis. Descriptive statistics such as means, frequencies and proportions were used to summarize the data. Bivariate analysis was done to determine associations between the predictor and outcome variables. Multivariable logistic regression was used to assess the strength of the association between the independent/ predictor variables and the outcome variables. A p-value of 0.05 was considered statistically significant. All results were summarized into tables and graphs.

### Quality control and assurance

The principal investigator recruited only RAs with a minimum of a bachelor’s degree in health sciences, social sciences, or humanities and any other related field. Prior to data collection, RAs were trained for two days on the data collection tool, ethics and study protocol. The data entry form was designed with skips and restrictions to ensure quality data collection. Pretesting of study tools was done at Kisenyi Health Centre IV by administering the data collection tool to pregnant women during their ANC visit in order to ensure familiarity of the RAs with the tool and also identify any errors. Also, to ensure compliance to the study protocol, RAs were supervised during data collection.

## Results

### Socio-demographic characteristics of participants

Of the 285 respondents, 41.0% (117/285) were aged between 18-24 years. The average age of the respondents was 26.6 years (SD ± 5.9). More than half, 59.2% (169/285) of the respondents were married/cohabiting, 6.7% (19/285) had no formal education, and 45.6% (130/285) had partners with tertiary level of education. About 46.0% (131/285) of the respondents were not employed, and 46.0% (131/285) had employed partners.

### Antenatal care attendance

More than half, 59.6% (170/285) of the respondents had knowledge of when to start ANC service, 59.3% (169/285) had received advice on when to start ANC and 61.4% (175/285) had an early first ANC visit (≤ 20 weeks). The average gravidity of respondents was 2.4 pregnancies (SD ± 1.8) and majority, 79.3% (226/285) had gravidity between 1-3 pregnancies. The average parity of respondents was 1.3 births (SD ± 1.7) and only 19.3% (55/285) had ≥3 births (Table 3).

**Table 1:** Socio-demographic characteristics of pregnant women attending ANC in Kawempe National Referral hospital.

| Variable | Frequency (N=285) | Percentage (%) |
| --- | --- | --- |
| **Mother's age group** |  |  |
| 18- 24 | 117 | 41 |
| 25-31 | 112 | 39.3 |
| 32-38 | 45 | 15.8 |
| 39-45 | 11 | 3.9 |
| **Marital status** |  |  |
| Married / cohabiting | 169 | 59.3 |
| Single/ not married | 85 | 29.8 |
| Divorced | 31 | 10.9 |
| **Mother's level of education** |  |  |
| More than secondary | 89 | 31.2 |
| No Education | 19 | 6.7 |
| Primary | 91 | 31.9 |
| Secondary | 86 | 30.2 |
| **Partner's level of education** |  |  |
| Above secondary | 130 | 45.6 |
| No Education | 15 | 5.3 |
| Secondary | 107 | 37.5 |
| Primary | 33 | 11.6 |
| **Respondent's employment status** |  |  |
| Employed | 95 | 33.3 |
| Not employed | 131 | 46 |
| Self employed | 59 | 20.7 |
| **Partner's employment status** |  |  |
| Employed | 131 | 46 |
| Self employed | 106 | 37.2 |
| Not employed | 48 | 16.8 |

**Table 2:**
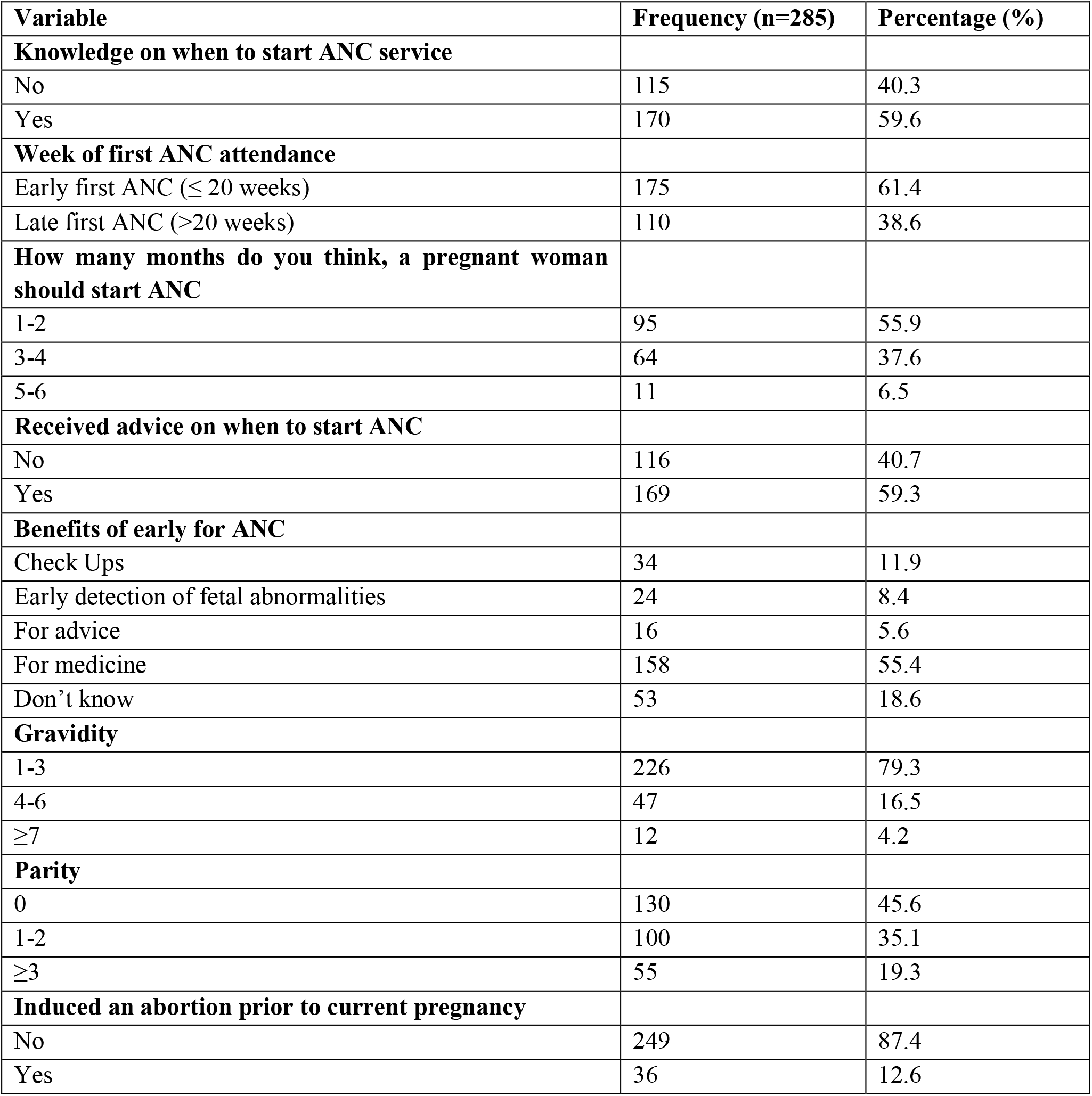
Antenatal care attendance and Obstetric factors among study participants.

**Table 3:**
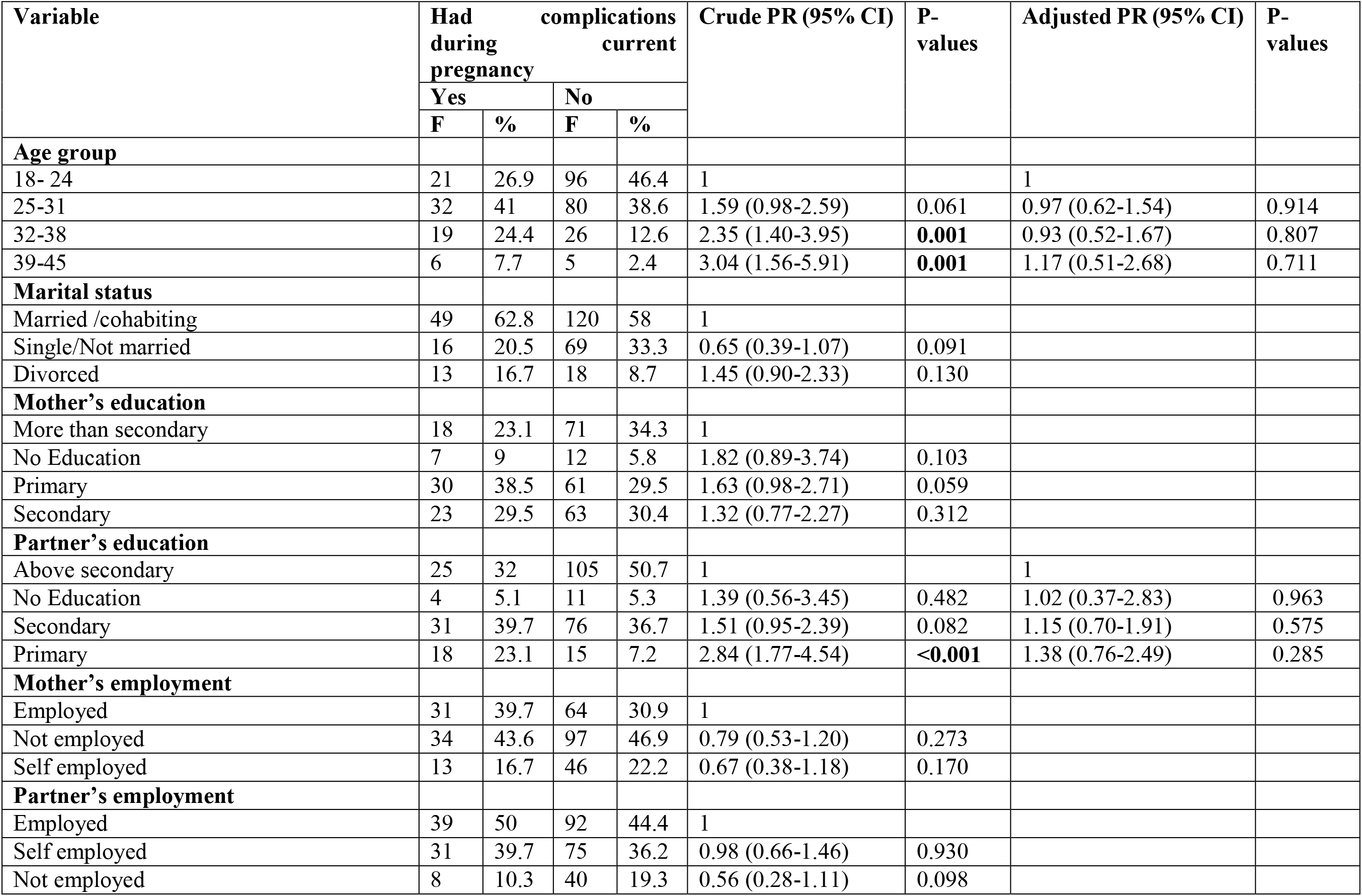
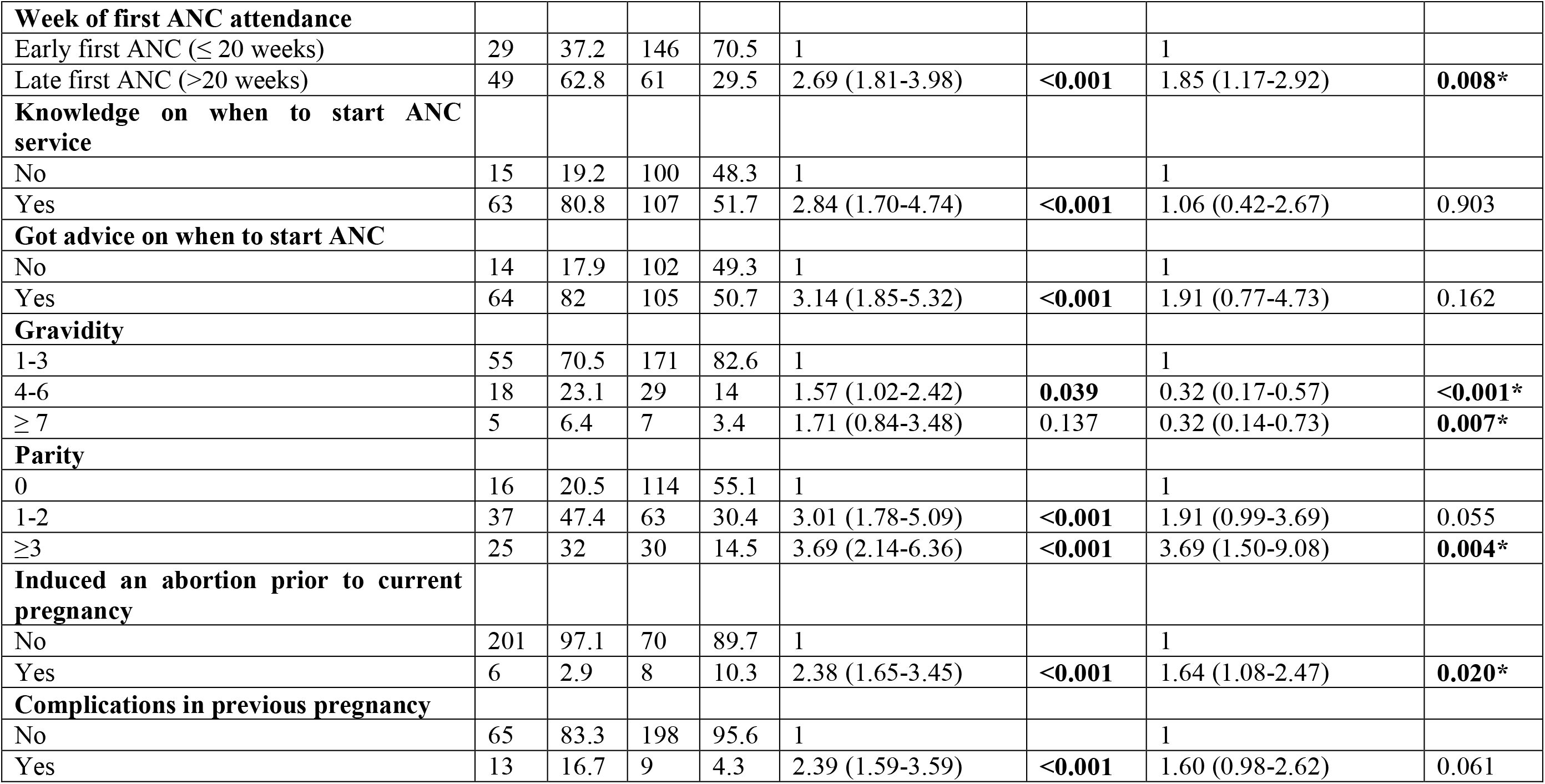
Factors associated with pregnancy-related complications among pregnant women attending ANC services at a specialised maternal and child health national referral hospital.

| Variable | Had complications during current pregnancy |  |  |  | Crude PR (95% CI) | P-values | Adjusted PR (95% CI) | P-values |
| --- | --- | --- | --- | --- | --- | --- | --- | --- |
|  | Yes |  | No |  |  |  |  |  |
|  | F | % | F | % |  |  |  |  |
| Age group |  |  |  |  |  |  |  |  |
| 18- 24 | 21 | 26.9 | 96 | 46.4 | 1 |  | 1 |  |
| 25-31 | 32 | 41 | 80 | 38.6 | 1.59 (0.98-2.59) | 0.061 | 0.97 (0.62-1.54) | 0.914 |
| 32-38 | 19 | 24.4 | 26 | 12.6 | 2.35 (1.40-3.95) | **0.001** | 0.93 (0.52-1.67) | 0.807 |
| 39-45 | 6 | 7.7 | 5 | 2.4 | 3.04 (1.56-5.91) | **0.001** | 1.17 (0.51-2.68) | 0.711 |
| Marital status |  |  |  |  |  |  |  |  |
| Married /cohabiting | 49 | 62.8 | 120 | 58 | 1 |  |  |  |
| Single/Not married | 16 | 20.5 | 69 | 33.3 | 0.65 (0.39-1.07) | 0.091 |  |  |
| Divorced | 13 | 16.7 | 18 | 8.7 | 1.45 (0.90-2.33) | 0.130 |  |  |
| Mother's education |  |  |  |  |  |  |  |  |
| More than secondary | 18 | 23.1 | 71 | 34.3 | 1 |  |  |  |
| No Education | 7 | 9 | 12 | 5.8 | 1.82 (0.89-3.74) | 0.103 |  |  |
| Primary | 30 | 38.5 | 61 | 29.5 | 1.63 (0.98-2.71) | 0.059 |  |  |
| Secondary | 23 | 29.5 | 63 | 30.4 | 1.32 (0.77-2.27) | 0.312 |  |  |
| Partner's education |  |  |  |  |  |  |  |  |
| Above secondary | 25 | 32 | 105 | 50.7 | 1 |  | 1 |  |
| No Education | 4 | 5.1 | 11 | 5.3 | 1.39 (0.56-3.45) | 0.482 | 1.02 (0.37-2.83) | 0.963 |
| Secondary | 31 | 39.7 | 76 | 36.7 | 1.51 (0.95-2.39) | 0.082 | 1.15 (0.70-1.91) | 0.575 |
| Primary | 18 | 23.1 | 15 | 7.2 | 2.84 (1.77-4.54) | **<0.001** | 1.38 (0.76-2.49) | 0.285 |
| Mother's employment |  |  |  |  |  |  |  |  |
| Employed | 31 | 39.7 | 64 | 30.9 | 1 |  |  |  |
| Not employed | 34 | 43.6 | 97 | 46.9 | 0.79 (0.53-1.20) | 0.273 |  |  |
| Self employed | 13 | 16.7 | 46 | 22.2 | 0.67 (0.38-1.18) | 0.170 |  |  |
| Partner's employment |  |  |  |  |  |  |  |  |
| Employed | 39 | 50 | 92 | 44.4 | 1 |  |  |  |
| Self employed | 31 | 39.7 | 75 | 36.2 | 0.98 (0.66-1.46) | 0.930 |  |  |
| Not employed | 8 | 10.3 | 40 | 19.3 | 0.56 (0.28-1.11) | 0.098 |  |  |
| **Week of first ANC attendance** |  |  |  |  |  |  |  |  |
| Early first ANC ($\leq 20$ weeks) | 29 | 37.2 | 146 | 70.5 | 1 | | 1 | |
| Late first ANC ($>20$ weeks) | 49 | 62.8 | 61 | 29.5 | 2.69 (1.81-3.98) | **<0.001** | 1.85 (1.17-2.92) | **0.008\*** |
| **Knowledge on when to start ANC service** |  |  |  |  |  |  |  |  |
| No | 15 | 19.2 | 100 | 48.3 | 1 |  | 1 |  |
| Yes | 63 | 80.8 | 107 | 51.7 | 2.84 (1.70-4.74) | **<0.001** | 1.06 (0.42-2.67) | 0.903 |
| **Got advice on when to start ANC** |  |  |  |  |  |  |  |  |
| No | 14 | 17.9 | 102 | 49.3 | 1 |  | 1 |  |
| Yes | 64 | 82 | 105 | 50.7 | 3.14 (1.85-5.32) | **<0.001** | 1.91 (0.77-4.73) | 0.162 |
| **Gravidity** |  |  |  |  |  |  |  |  |
| 1-3 | 55 | 70.5 | 171 | 82.6 | 1 |  | 1 |  |
| 4-6 | 18 | 23.1 | 29 | 14 | 1.57 (1.02-2.42) | **0.039** | 0.32 (0.17-0.57) | **<0.001\*** |
| $\geq 7$ | 5 | 6.4 | 7 | 3.4 | 1.71 (0.84-3.48) | 0.137 | 0.32 (0.14-0.73) | **0.007\*** |
| **Parity** |  |  |  |  |  |  |  |  |
| 0 | 16 | 20.5 | 114 | 55.1 | 1 |  | 1 |  |
| 1-2 | 37 | 47.4 | 63 | 30.4 | 3.01 (1.78-5.09) | **<0.001** | 1.91 (0.99-3.69) | 0.055 |
| $\geq 3$ | 25 | 32 | 30 | 14.5 | 3.69 (2.14-6.36) | **<0.001** | 3.69 (1.50-9.08) | **0.004\*** |
| **Induced an abortion prior to current pregnancy** |  |  |  |  |  |  |  |  |
| No | 201 | 97.1 | 70 | 89.7 | 1 |  | 1 |  |
| Yes | 6 | 2.9 | 8 | 10.3 | 2.38 (1.65-3.45) | **<0.001** | 1.64 (1.08-2.47) | **0.020\*** |
| **Complications in previous pregnancy** |  |  |  |  |  |  |  |  |
| No | 65 | 83.3 | 198 | 95.6 | 1 |  | 1 |  |
| Yes | 13 | 16.7 | 9 | 4.3 | 2.39 (1.59-3.59) | **<0.001** | 1.60 (0.98-2.62) | 0.061 |

### Pregnancy-related complications

Out of the 285 women, 27.4% (78/285) had complications in current pregnancy. The prevalence of complications related to the current pregnancy was 10.9% (31/285) for anaemia; 8.1% (23/285) for eclampsia; 4.9% (14/285) for still births; 2.1% (6/285) for gestational diabetes; 1.4% (4/285) for miscarriage; and 0.7% (2/285) for pre-eclampsia (Fig 1). Additionally, only 7.7% (22/285) of the respondents had complications in preceding pregnancy. The prevalence of complications in the preceding pregnancy was 2.8% (8/285) for eclampsia; 1.0% (3/285) for postpartum haemorrhage; 0.7% (2/285) for antepartum haemorrhage; 0.7% (2/285) for anaemia; 0.7% (2/285) for malaria; 0.7% (2/285) for placenta previa; 0.3% (1/285) for gestational diabetes; 0.3% (1/285) for painful fibroids; and 0.3% (1/285) for UTIs.

**Figure 1:**
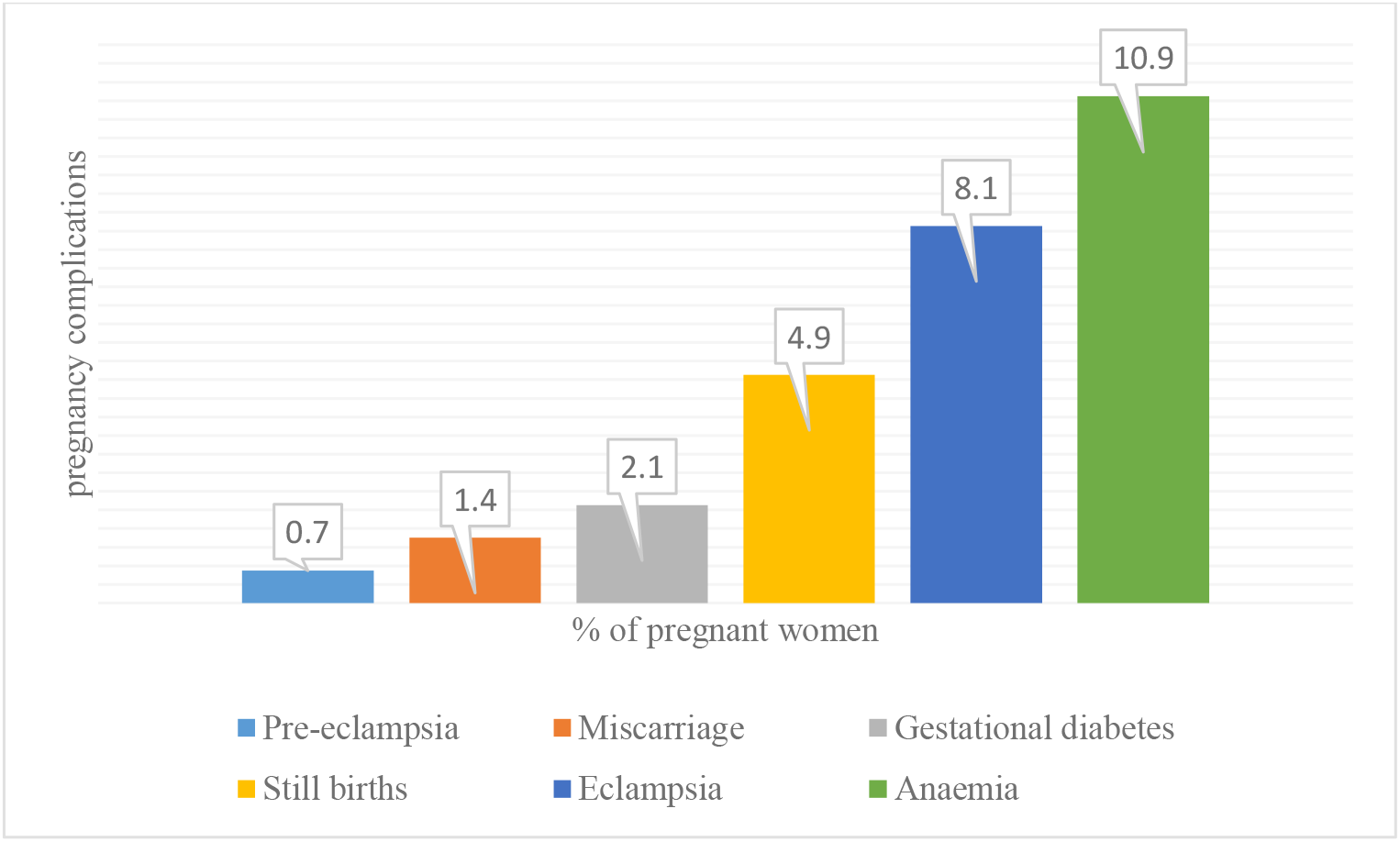
Complications among pregnant women attending ANC at Kawempe national referral hospital

### Factors associated with pregnancy-related complications among women attending ANC at a specialised maternal and child health national referral hospital

At bivariate analysis, respondents were more likely to have pregnancy-related complications if they were age between 32-38 years and aged between 39-45 years as compared to those aged between 18-24 years. Respondents whose partners had primary education had a 184% higher likelihood of having pregnancy-related complications as compared to those whose partners had tertiary education (CPR= 2.84, 95% CI: 1.77-4.54). Also, respondents who had a late first ANC (>20 weeks) had a 169% higher likelihood of having pregnancy-related complications as compared to those who had an early first ANC visit (≤20 weeks) (CPR= 2.69, 95% CI: 1.81-3.98). Respondents who had knowledge on when to start ANC services had a 184% higher likelihood of having pregnancy-related complications as compared to those who didn’t have (CPR= 2.84, 95% CI: 1.70-4.74).

Additionally, respondents who indicated to have got advice on when to start ANC had a 114% higher likelihood of having pregnancy-related complications as compared to those who reported otherwise (CPR= 3.14, 95% CI: 1.85-5.32). Respondents with gravidity between 4-6 pregnancies had a 57% higher likelihood of having pregnancy-related complications as compared to those with gravidity between 1-3 pregnancies (CPR= 1.57, 95% CI: 1.02-2.42). Respondents who had a parity between 1-2 births (CPR= 3.01, 95% CI: 1.78-5.09) and those who had a parity of ≥3 births (CPR= 3.69, 95% CI: 2.14-6.36) were more likely to have pregnancy-related complications as compared to those who had no births yet. Having induced an abortion prior to current pregnancy (PR= 2.38, 95% CI: 1.65-3.45) and having complications in previous pregnancy (CPR= 2.39, 95% CI: 1.59-3.59) were positively associated with having pregnancy-related complications.

After adjusting for potential confounders, respondents who had a late first ANC (>20 weeks) had an 85% higher likelihood of having pregnancy-related complications as compared to those who had an early first ANC visit (≤20 weeks) (APR= 1.85, 95% CI:1.17-2.92). Respondents who had a gravidity between 4-6 pregnancies had a 32% lower likelihood of having pregnancy-related complications as compared to those who had gravidity between 1-3 pregnancies (APR= 0.32, 95% CI: 0.17-0.57). Also, Respondents who revealed to have a gravidity greater than 7 pregnancies had a 32% lower likelihood of having pregnancy-related complications as compared to those who had gravidity between 1-3 pregnancies (APR= 0.32, 95% CI: 0.14-0.73). Moreover, respondents who had a parity of ≥ 3 births had a 269% higher likelihood of having pregnancy-related complications as compared to those who had never given birth yet (APR= 3.69, 95% CI: 1.50-9.08). Having had an abortion prior to current pregnancy was positively associated with pregnancy-related complications (APR= 1.64, 95% CI: (1.08-2.47).

## Discussion

This study established pregnancy-related complications and associated factors among women attending ANC at a specialised maternal and child health national referral hospital in Uganda. More than quarter of the respondents had pregnancy-related complications. The most reported complications were anaemia, eclampsia and still births. Having a late first ANC (>20 weeks), a gravidity greater than 4, a parity greater than 3, and history of an induced abortion prior to the current pregnancy were associated with having pregnancy-related complications.

The prevalence of pregnancy-related complications (27.4%) among women attending ANC was very high. The high prevalence of pregnancy-related complications among women attending ANC at the study healthcare facility is not surprising, given the fact that it is a specialised maternal and child health national referral hospital (21). Such a healthcare facility receives referrals of more complicated cases which may not be managed at lower level or other specialised healthcare facilities. The prevalence of pregnancy-related complications is higher than that reported in Ethiopia (15.9%) (22) and Rwanda (13.8%) (23). The variations in the prevalence may have resulted from differences in the study context, population and designs used. The study in Ethiopia was community-based and retrospective (22), while that in Rwanda was cross-sectional in nature and involved lower level healthcare facilities (23). Our study was done in a specialised maternal and child health national referral hospital. Our study, just like others (23-27), reported anaemia, eclampsia, and still births as the most prevalent complications. The high prevalence of pregnancy-related complications reported in our study may be attributed to poor pregnancy planning and limited access to information regarding a healthy pregnancy (28-32).

We found out that anaemia was the most prevalent complication (10.9%). The prevalence of anaemia in the current study is however, lower than that reported in two regional referral hospitals in Uganda (22%) (33) and that reported by Bongomin, Olum (34) in Kawempe specialised national referral hospital (14.1%). The difference in the prevalence may be attributed to the difference in the study design and measurement of anaemia. Our study used self-reports (which are subject to social desirability bias) while Bongomin, Olum (34) used mixed designs (a cross-sectional, systematic review and meta-analysis), and assessed anaemia using the HumaCount 5D Haematology System. The high prevalence of anaemia reported in the current study may be explained by the high parity and gravidity among mothers in our study. About 19.3% of the women in our study has a parity of greater than three and more than 20.7% had a gravidity greater than 4. There is evidence that a high gravidity and/or is associated with pregnancy-related complications (35-38). Other factors known to cause iron deficiency anaemia are a lack of a diet rich in iron, infections, and genetics (33). Besides, anaemia, eclampsia was also highly prevalent. The high prevalence of eclampsia is worrying, given that it is one of the leading causes of maternal mortality (39). Eclampsia if not well managed, can lead to fetal growth restriction, preterm birth, placental abruption leading to haemorrhage, and death (40-43).

More than a third of the women in our study had a late ANC attendance. Late ANC attendance could be attributed to structural barriers such as lack of transport, poverty, limited information on pregnancy care, and un supportive spouses and communities (44). Women who received their first ANC after 20 weeks had a higher likelihood of having pregnancy-related complications as compared to those who received ANC before 20 weeks. Early ANC attendance facilitates the identification of risk factors for poor pregnancy or childbirth outcomes. In Uganda, the first ANC provides an opportunity for health education and promotion on pregnancy care, common pregnancy complaints, nutrition, danger signs, hygiene, rest and exercise. It is at this point that vital examinations such as blood pressure, pulse, and tests on HIV, syphilis test and hepatitis B (15). Women who are unable to attend ANC clinics at this point therefore, cannot benefit from these services, yet they are known to contribute to pregnancy-related complications (45-48). The role of early ANC in improving pregnancy outcomes has also been documented in previous studies (49-51). Our finding suggests the need to create awareness of the role of early ANC attendance in reducing pregnancy-related complications.

Having a parity greater than three increased the likelihood of having pregnancy-related complications as compared to those who had never given birth. There is evidence that an increase in the number of births increases the risk of pregnancy-related complications. Giving birth too many times is often associated with placenta previa, a condition known to increase the risk of pregnancy-related complications (52). Indeed, a systematic and modelling analysis by Chawanpaiboon, Vogel (53) revealed that women who had given births more than five times had an increased risk of giving birth to preterm infants. There is also evidence that parity increases the risk of anaemia, diabetes mellitus, eclampsia, malpresentation, abruption placentae, placenta previa, post-partum haemorrhage due uterine atony, and uterine rupture (54-59). Findings from this study reveal the need to sensitise mothers on the dangers of giving birth to many children, and to increase access to family planning education and services.

The likelihood of pregnancy-related complications reduced with an increase in gravidity. This reduction may be attributed to increasing experience in managing pregnancy. Women who have carried pregnancies more often are more likely to be aware of how to prevent pregnancy-related complications compared to those who have not. Our finding is consistent with that of Andemel, Gaoussou (60) which indicated that a low gravidity was associated with preterm birth. However, our findings also deviate from those of earlier studies, which indicated that an increase in gravidity increased the likelihood of pregnancy-related complications (61). Aside gravidity, having had an induced abortion prior to current pregnancy was positively associated with pregnancy-related complications. Abortion has been linked to complications such as damage to the womb or cervix, excessive bleeding, infection of the uterus or fallopian tubes, scarring of the inside of the uterus, sepsis or septic shock and uterine perforation (62, 63). Although these complications may be short term, studies have reported long term side effects of abortion such as weakening of the cervix which predisposes women to pregnancy-related complications and even preterm deliveries (64, 65). For the case of Uganda, abortion is illegal unless performed by a licensed medical doctor to save a woman’s life. With women lacking safe and legal options for abortion and a high rate of unintended pregnancies, many resort to unsafe and high-risk abortion practices such as self-induced abortions (66, 67). There’s therefore a need to legalize abortion so that women can access safe abortion services thus reducing pregnancy-related complications. Also, interventions aimed at controlling unintended pregnancies such as improving awareness and access to on family planning services.

## Strengths and limitations

This is one of the few studies that has so far explored the prevalence of pregnancy-related complications among women attending antenatal care in a specialised maternal and child health national referral hospital in a low-resource setting. It used a relatively large sample size which is representative. However, our study had the limitation of using self-reports, which are subject to social desirability bias (68).

## Conclusion

The prevalence of pregnancy-related complications (27.4%) among women attending ANC was very high. The most reported pregnancy-related complications were anaemia, eclampsia and still births. Having a late first ANC (>20 weeks), a gravidity greater than 4, a parity greater than 3, and history of an induced abortion prior to the current pregnancy were associated with having pregnancy-related complications. Therefore, interventions aimed at promoting early ANC attendance, increasing access to family planning education and services, and increasing access to safe abortion services are vital in the reduction of pregnancy-related complications.

## Data Availability

The datasets used and/or analysed during the current study are not publicly available, but are available from the corresponding author on reasonable request.

## Acknowledgement

Gratitude goes to Kawempe national referral hospital administration for permitting us to conduct the study and for the technical support provided. Credit goes research assistants for collecting data with due diligence

## Abbreviations

ANC: Antenatal Care
MOH: Ministry of Health
RAs: Research Assistants
SSA: Sub Saharan Africa
UTIs: Urinary Tract Infections
WHO: World Health Organization

## Ethical declarations

Ethical approval was sought from Makerere University School of Health Sciences Research and Ethics Committee and permission to conduct this study was gotten from the medical director Kawempe National Referral hospital. All participants were thoroughly informed about the study and were required to give informed consent prior to participation. Confidentiality and privacy was highly observed, no name of the participant appeared in the questionnaire, as only the participant was required to fill the questionnaire.

## Author contributions

BNT, DB, AVJ and TS participated in study conceptualisation. BNT, DB, JBI, AVJ, JBN, AT, AN, RM, RKM, TS and DL participated in the analysis and drafting of the manuscript. All authors reviewed and approved the final manuscript.

